# Updated Model for the USA Summer 2021 CoVID-19 Resurgence

**DOI:** 10.1101/2021.10.15.21265078

**Authors:** Genghmun Eng

## Abstract

Over the course of the CoVID-19 pandemic, we utilized widely-available real-time data to create models for predicting its spread, and to estimate the time evolution for each of the USA CoVID-19 waves. Our recent *medrxiv*.*org* preprint (*10*.*1101_2021*.*08*.*16*.*21262150*) examined the USA Summer 2021 resurgence, from ∼6/7/2021 up through ∼8/15/2021 (*Stage 1*). Our preprint covering this period showed that CoVID-19 could infect virtually all susceptible non-vaccinated persons, who were practicing minimal *Social Distancing* and NO *Mask-Wearing*.

The most recent USA Summer 2021 resurgence data, from ∼8/13/2021 up through 10/7/2021 (*Stage 2*), shows a significant “flattening of the curve”. Since no new government mandates were involved, our interpretation is that some vaccine-hesitant people have now elected to become vaccinated. The *Social Distancing* parameter in our model showed a ∼6.67X increase between *Stage 1* and *Stage 2*, indicating that this parameter also can serve as an indicator of vaccination rates. The other parameter in our model, which is associated with *Mask-Wearing*, increased from zero to a finite but relatively small value. Using the 10/7/2021 USA CoVID-19 overall mortality rate of ∼1.60942 % gives these updated predictions for the total number of USA CoVID-19 cases and deaths:

*N*_*TOTAL*_(3*/*21*/*2022) ≈ 52, 188, 000 ; *N*_*Deaths*_(3*/*21*/*2022) ≈ 839, 900, *N*_*TOTAL*_(3*/*21*/*2024) ≈ [52, 787, 000 ; *N*_*Deaths*_(3*/*21*/*2024) ≈ 849, 600, assuming no new 2021 Winter Resurgence occurs (*with 3 Figures*).

## 1 Introduction

Each new wave of the USA CoVID-19 pandemic started with a sharp rise in the total number *N*(*t*) of new cases above the prior baseline. Since the exact start of each wave is not easily determined, the *t* = 0 point for each wave was chosen to be when the resurgence was easily identified, where *N*(*t* = 0) = *N*_*o*_ is the number of cases above baseline that have already occurred.

In March 2020, California Governor Gavin Newsom began the first large-scale efforts at pandemic mitigation, mandating state-wide lockdowns, “stay at home” orders, school closures, restrictions on business operations, and new *Social Distancing* requirements, including minimum separation distances and decreased allowable occupancy. Other states soon followed.

Since then, whenever the pandemic appeared to be beaten down, restrictions were relaxed, CoVID-19 cases increased, new CoVID-19 variants appeared, and the pandemic rose up again, almost with every season. The daily number of new cases *dN*(*t*) */ dt* clearly shows the initial Spring 2020 pandemic start, a Summer 2020 resurgence, and a long and pernicious Winter 2020 “*Third Wave*”. A *dN*(*t*) */ dt* uptick in Spring 2021 was actually a small “*Fourth Wave*”, which would make the *present-day* Summer 2021 resurgence a “*Fifth Wave*”. However, most CoVID-19 reporting sites ignore the Spring 2021 uptick, labeling the entire Summer 2021 resurgence as a “*USA Fourth Wave*”.

Our mathematical modeling^**1-5**^ is empirically based, and it does not predict when each new CoVID-19 wave will start, or what biological and social circumstances are causing the wave. However, once the wave has begun, our models can successfully predict its time evolution.

The standard **SEIR** (Susceptible, **E**xposed, **I**nfected, and **R**ecovered or Removed) epidemiology model starts with this basic function for exponential growth (*R*_*o*_ *>* 1) or decay (*R*_*o*_ *<* 1):

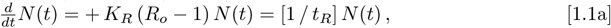

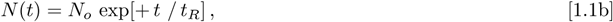

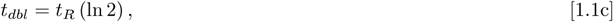

where *t*_*R*_ *>* 0 is the pandemic growth rate, and *t*_*dbl*_ is the doubling-time for *N*(*t*). Given a total population of *N*_*ALL*_, the *uninfected population* is:

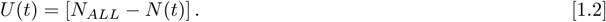

Using Eq. [1.1a] implicitly assumes that *N*_*ALL*_ is large enough so that *N*(*t*) *<< N*_*ALL*_ for all times of interest, so that pandemic saturation effects do not need to be separately considered.

Much of the **SEIR** modeling examines the major factors that are involved in setting the {*K*_*R*_, *R*_*o*_, *t*_*R*_} values. The {*K*_*R*_, *R*_*o*_, *t*_*R*_} parameters quantify how much contact the infected *N*(*t*) group has with the *uninfected population*, with those contacts determining the next *N*(*t* + Δ*t*) value. This feature makes Eq. [1.1a] a local transmission model.

As such, the **SEIR** models do not explicitly consider what the *U*(*t*) *uninfected population*, as a whole, may be doing in response to the pandemic, prior to becoming infected. Factors such as *Social Distancing, Mask-Wearing, Vaccination*, and large-scale *Government Mandates*, each can affect *U*(*t*), which then introduces new *non-local factors* into the pandemic evolution.

Following our original CoVID-19 preprint^**1-2**^ of April-May 2020, the Eqs. [1.1a]-[1.1c] **SEIR** Model for pandemic evolution was first extended by introducing a new parameter {*α*_*S*_} to account for new *Social Distancing* practices among the *uninfected population*:

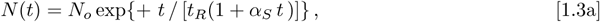

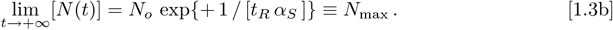

The resulting Eq. [1.3b] shows that any non-zero *Social Distancing α* _*S*_ value results in a natural pandemic end, prior to infecting the entire *N*_*ALL*_ population. In our follow-on pre-prints^**3-5**^, this basic model was extended to handle the case when the [*dN*_*data*_(*t*) */ dt*] data showed a post-peak exponential decay with time:

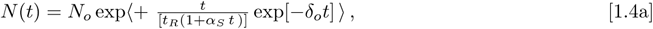

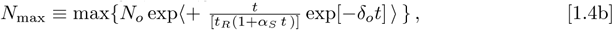

instead of the [1*/t*^*2*^] dependence predicted by Eq. [1.3a]. This modification more accurately predicts when the 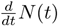 peak occurs, and gives an exponential decay “tail” for the daily number of new cases.

However, post-peak, when the 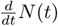 values become small, Eq. [1.4a] can reach a point where 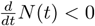 first occurs. Although it pinpoints a particular time for the pandemic end, this value is likely inaccurate. Since 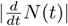 is small at that point, it will usually be outside the domain of interest, as the actual pandemic will be nearly over by then, barring a follow-on resurgence.

## 2 Updated Summer 2021 Resurgence Projections

Since we have successfully modeled the various prior USA CoVID-19 waves^**1-5**^ using the same few parameters, that modeling success indicates that the **response** of the *U*(*t*) *uninfected population* has been similar for each CoVID-19 wave, even if different factors were driving each new resurgence.

The latest USA Summer 2021 CoVID-19 wave follows this trend. Our 8/15/2021 preprint^**5**^ showed that this latest wave started around ∼6/7/2021. Hospital admission records indicated that unvaccinated persons primarily comprised this *N*(*t*) infected group. Interviews with those CoVID-19 patients found that many were resistant to vaccination, even when it was available.

Our modeling up through 8/15/2021 showed that the CoVID-19 spread among this group was associated with very little *Social Distancing* and virtually NO *Mask-Wearing*. As given in our prior pre-print^**5**^, the parameter values for this initial portion (“*Stage 1* “) of USA Summer 2021 resurgence were:

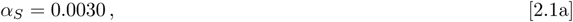

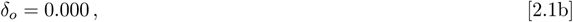

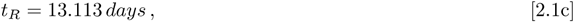

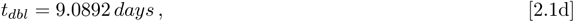

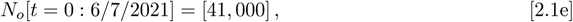

for the period of 6/7/2021 to 8/15/2021. The *N*_*o*_[*t* = 0] value in Eq. [2.1e] shows how many resurgence cases above baseline were needed to identify that a resurgence had started, rather than being a data reporting fluctuation. These {*α*_*S*_; *δ*_*o*_} parameter values were so small that virtually every susceptible non-vaccinated USA person could potentially become infected, due to their “minimal *Social Distancing* “ and “NO Mask-Wearing” practices.

The *N*(*t*) data from the next two months shows that the situation has markedly improved. The new *N*(*t*) shape changes were similar to how the original March 2020 mandates changed the course of the CoVID-19 pandemic, with the *N*(*t*) function for this wave now showing a substantial “flattening of the curve” between mid-August 2021 and the *present-day* (10/7/2021), meriting this update.

Another commonality that appears to exist among the various USA CoVID-19 waves is that the *U*(*t*) *unifected population* group seems to ignore the pandemic resurgence until it reaches a level where the local hospitals begin to fill up with CoVID-19 patients, and the local media fills up with their dire stories.

Updating the USA Summer 2021 resurgence with data from ∼8/13/2021 through 10/7/2021, this latter (“*Stage 2* “) portion of the USA Summer 2021 resurgence now gives these new parameter values:

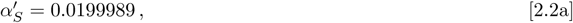

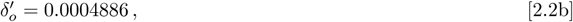

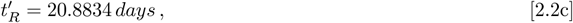

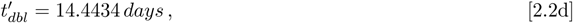

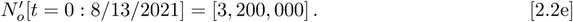

The large 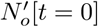 value in Eq. [2.2e] likely indicates how many resurgence cases above baseline were needed before the *uninfected population* began, as a group, to alter their average behavior.

The combined *Stage 1* and *Stage 2* predictions are graphically illustrated in *Figure 1* covering the entire USA Summer 2021 CoVID-19 resurgence by itself, from ∼6/7/2021 up through 10/7/2021, as all prior CoVID-19 waves were first subtracted out as a baseline. A good datafit between the *N*(*t*) predictions and the raw *N*_*data*_(*t*) values is evident, along with a clear “curve flattening” for *Stage 2* of this CoVID-19 wave.

**Fig. 1:**
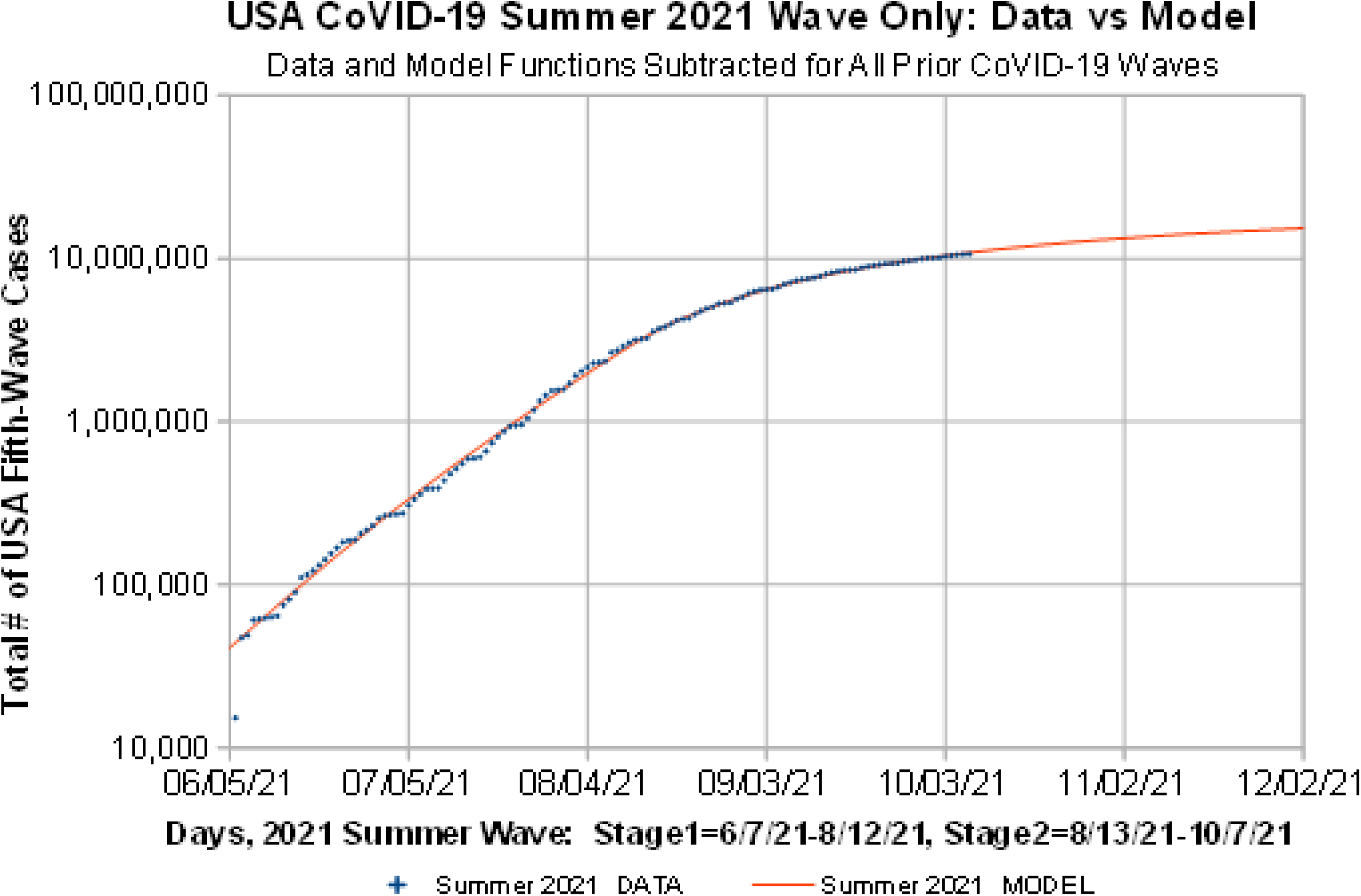
The USA CoVID-19 Summer 2021 Resurgence, By Itself.

These *Figure 1* USA Summer 2021 resurgence values were then added to the model baseline data for all the previous USA CoVID-19 waves. Those results, shown in *Figure 2*, indicate that we are presently in the 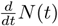 post-peak phase, with the rather small 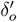 value of Eq. [2.2b] giving the gradually decreasing longtail in *Figure 2*, for the predicted USA 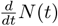 future progression.

**Fig. 2:**
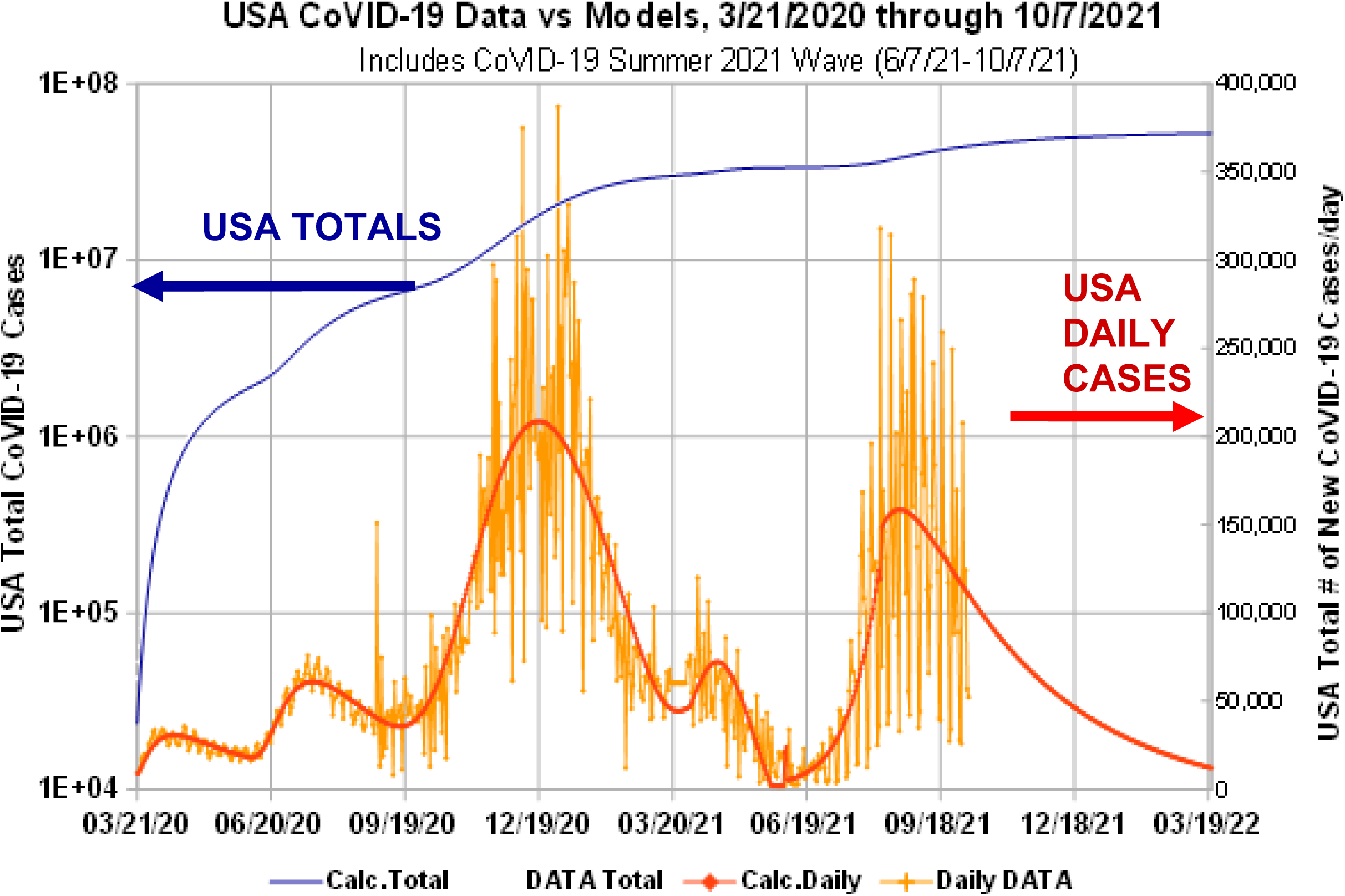
USA CoVID-19 Totals: 3/21/2020 through 10/7/2021.

A comparison of these new 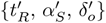 values and the {*t*_*R*,_ *α*_*s*,_*δ*_*o*_} values from all the previous USA CoVID-19 waves is shown in *Figure 3*. The Eq. [2.2a] *Social Distancing* parameter 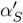 for *Stage 2* is about ∼6.67 *X* larger than the *α*_*S*_ value of Eq. [2.1a] for *Stage 1*. The 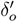 value for *Stage 2*, which is associated with *Mask-Wearing*, is still quite small, being ∼3.58*X* smaller than the least of the previous non-zero *δ*_*o*_-values. Additional *Mask-Wearing* by this substantially unvaccinated group would help end this pandemic wave faster.

**Fig. 3:**
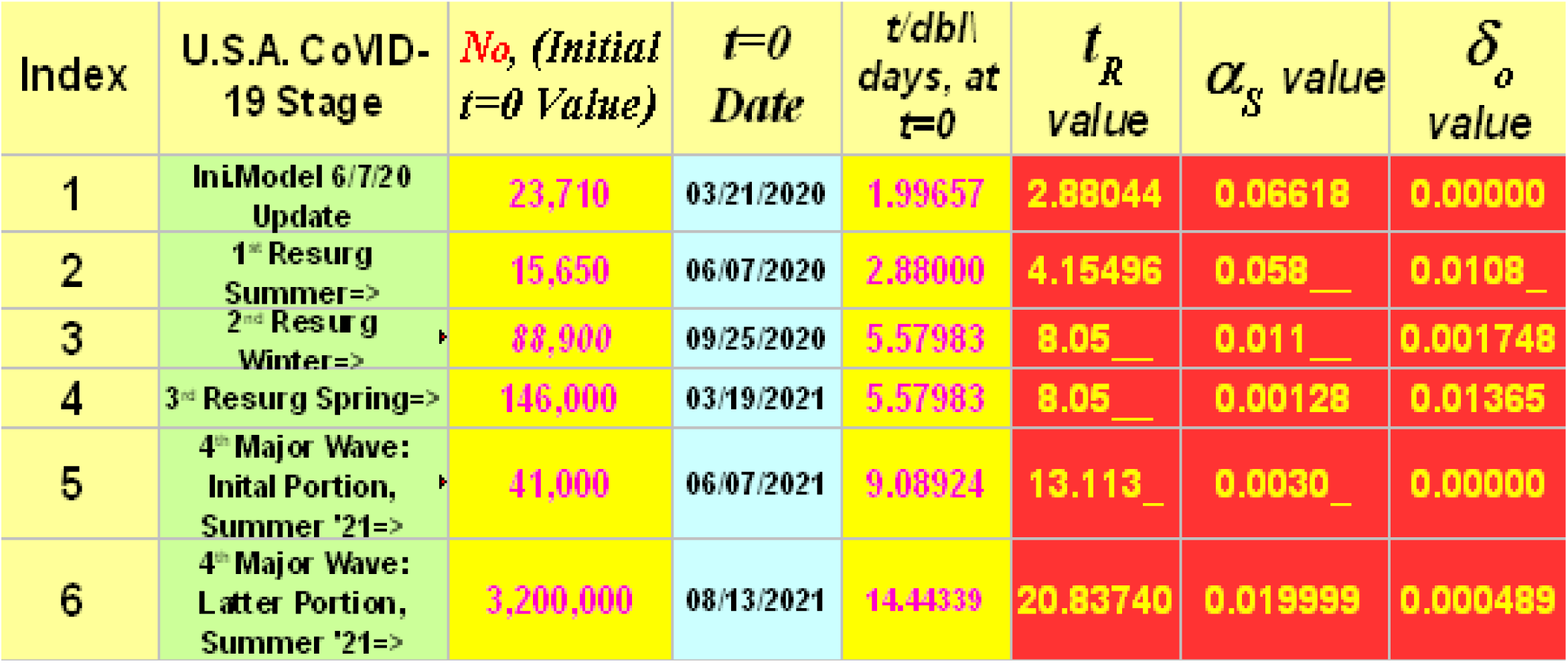
Summary of CoVID-19 Models and Parameter Values.

The change in the value from *α*_*S*_ to 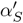 is good, since it occurred without any significant new government mandates being put in place. Our best interpretation for what changed is that more of the previously vaccine-hesitant group have now become vaccinated. Thus, *α*_*S*_ and 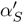 likely now measure vaccination effects as the primary new *Social Distancing* parameter.

Using Eq. [2.2a]-[2.2c], the USA Summer 2021 resurgence, by itself, gives a total number of new CoVID-19 resurgence cases:

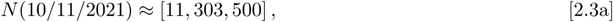

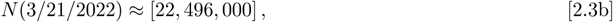

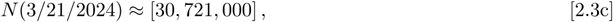

above the prior baseline. Adding these new resurgence-only results to our values for the prior USA CoVID-19 waves gives these updated predictions:

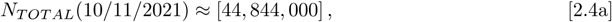

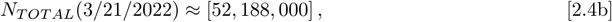

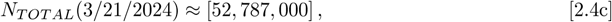

for the total number of USA CoVID-19 cases. Using the latest overall USA CoVID-19 mortality rate (10/7/2021):

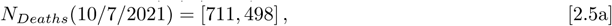

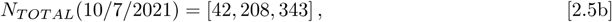

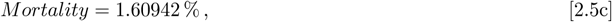

then sets these future mortality estimates:

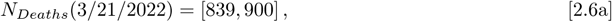

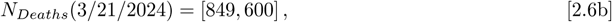

assuming no new 2021 Winter Resurgence occurs.

## 3 Summary

The USA Summer 2021 resurgence, which began around ∼6/7/2021, persists to the *present-day* (∼10/7/2021). Our initial model for this CoVID-19 wave (“*Stage 1* “), up through ∼8/15/2021, as given in our prior pre-print^**5**^, identified this CoVID-19 wave as being primarily among the unvaccinated. Our analysis also showed that this subgroup was practicing very little *Social Distancing*, with virtually NO Mask-Wearing.

The most recent data for this CoVID-19 wave (“*Stage 2* “), as analyzed here, shows that the period of ∼8/13/2021 through the *present-day*, has a significant “flattening of the curve”, as was shown in Figure 1. The new 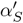 *Social Distancing* parameter for this *Stage 2* of the USA Summer 2021 resurgence is about 6.67 *X* larger than the *α*_*S*_ value for the prior *Stage 1*. As this change in the *Social Distancing* parameter value was not associated with any new government mandates, this change is likely due to increased vaccination among the CoVID-19 susceptible population. Thus, the *α*_*S*_ model parameter in Eq. [1.3a] may now track vaccination rates.

However, as shown in Figure 2, the *present-day* 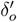 value, which is associated with *Mask-Wearing*, is one of the smallest non-zero values found so far, for any USA CoVID-19 wave. It is ∼3.58*X* smaller than all prior non-zero *δ*_*o*_-values, indicating that more *Mask-Wearing* by this substantially unvaccinated group is also warranted.

These parameter values give these updated predictions:

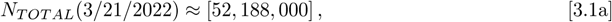

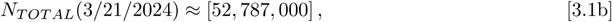

for the total number of USA CoVID-19 cases. The *present-day* USA CoVID-19 mortality rate of 1.6094 %, gives these future mortality estimates:

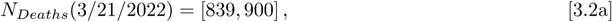

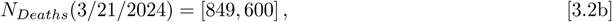

assuming no new 2021 Winter Resurgence occurs.

## Data Availability

All data produced in the present work are contained in the manuscript.

